# cocci-call: a species-aware variant identification pipeline for *Coccidioides* spp. in genomic epidemiology

**DOI:** 10.1101/2025.01.14.25320518

**Authors:** Marco Marchetti, Emanuel M. Fonseca, Kimberly E. Hanson, Bridget Barker, Katharine S. Walter

**Author notes:** **CORRESPONDENCE:** Katharine S. Walter. **REPOSITORIES:** Raw sequencing data and the cocci-call pipeline, along with all analysis scripts, are publicly available under BioSample SAMN45887705 and at https://github.com/ksw9/cocci-call.

## Abstract

Emerging fungal pathogens, such as *Coccidioides*, the causative agent of Valley fever, pose significant clinical and public health challenges. While advances in genomic epidemiology have enhanced our understanding of *Coccidioides* evolutionary history, the lack of standardized tools for variant identification makes it difficult to draw comparisons between studies. To address this gap, we developed and benchmarked a novel, publicly available pipeline, cocci-call, designed for genome-wide variant identification and species assignment. We found that cocci-call correctly identifies *Coccidioides* species in 100% (20/20) *in silico* sequence read sets and 98.86% (173/175) clinical or environmental samples. We found that performance of variant identification significantly improved when focusing on high-confidence genomic benchmarking regions. For example, the F1 score increased from 63.9% to 93.8% for *C. posadasii* and 60.2% to 84.7% *C. immitis*, respectively, when moving from a whole-genome analysis to gene-only analysis. We observed a similar trend when repetitive regions were masked. Empirical sequencing of the *C. posadasii* NR-166 reference strain corroborated these findings, with cocci-call achieving an F1 score of 63.9% in whole-genome analysis and 93.8% in gene-only regions. These results underscore the robustness and utility of cocci-call for *Coccidioides* genomic analyses, making it a valuable tool for genome-wide association studies and phylogenetics of *Coccidioides*.

**DATA SUMMARY:** Raw sequencing data generated for this study are available in the Sequence Read Archive (SRA) under BioSample SAMN45887705. This includes whole-genome sequencing data used for variant identification and benchmarking, as well as sequencing data from clinical and environmental Coccidioides samples. The pipeline cocci-call, developed for species-aware variant identification in *Coccidioides* spp., is publicly available at https://github.com/ksw9/cocci-call. All scripts and code used for the analysis performed in this study are included in the pipeline repository.

**IMPACT STATEMENT:** Coccidioidomycosis or Valley fever, caused by the fungal pathogens *Coccidioides* spp., is a growing public health concern. Understanding the genetic diversity of these pathogens is vital for tracking infections, improving diagnostics, and guiding effective treatments. Despite its public health importance, the lack of standardized tools for analyzing *Coccidioides* genomes has hindered the comparability of findings across studies. To address this challenge, we developed and measured the performance of cocci-call, an open-source pipeline for species assignment and variant identification. By establishing a standardized approach, cocci-call facilitates cross-study comparisons, enhances collaboration among research groups, and contributes to a deeper understanding of pathogen evolution and transmission dynamics. Ultimately, our study supports global efforts to combat fungal infections and protect public health by providing a consistent, benchmarked framework for genomic research.

## INTRODUCTION

Emerging fungal pathogens are an escalating public health threat, demanding urgent attention from the global health community (1). In 2022, the World Health Organization (WHO) underscored this urgency by publishing its first-ever list of fungal pathogens prioritized for public health research, placing *Coccidioides* spp., the causative agents of coccidioidomycosis or Valley Fever, among the most critical (2). Valley Fever is caused by two closely related fungal species, *Coccidioides immitis* and *C. posadasii*, which exhibit distinct geographical distributions. *Coccidioides immitis* predominantly occurs in California, Baja California (Mexico), and Washington state, while *C. posadasii* is more widespread, spanning Arizona, New Mexico, Texas, and various regions of Latin America (3). Despite its substantial health implications, coccidioidomycosis remains underdiagnosed, complicating disease management and hindering accurate assessments of its impact on healthcare systems (4). Given *Coccidioides* spp. ability to cause both local and travel-associated infections, there is an urgent need for advanced diagnostic tools that enable rapid and accurate identification, allowing for better disease tracking, and improved patient management (5,6).

However, the full potential of advanced genomic tools, such as whole-genome sequencing (WGS), has yet to be fully realized for *Coccidioides* spp. due to the lack of standardized bioinformatics workflows. Genomic studies face challenges from the absence of well-validated pipelines for variant identification and tools for accurate species-level assignment, both of which are essential for better tracking and understanding the disease. Current workflows are often customized, making it difficult to compare results across studies and limiting the broader application of genomic data. Addressing these gaps is crucial—not only for *Coccidioides* spp. but also for other fungal pathogens. For example, efforts to standardize variant detection in *Candida auris* studies have significantly improved reliability and consistency (7). Developing standardized bioinformatics tools tailored to the unique challenges of *Coccidioides* spp. will help advance genomic research and improve the management of coccidioidomycosis.

In response to these challenges, we developed and benchmarked a species-aware variant identification pipeline, cocci-call, tailored specifically for *Coccidioides* spp. Our pipeline detects genome-wide single nucleotide polymorphism (SNP) variants and assigns species, supporting future genomic epidemiology and association studies. We assessed the performance of cocci-call alongside another fungal variant identification tool, MycoSNP (7), using both simulated and empirically generated sequencing data. Additionally, we tested cocci-call’s accuracy in assigning species in a collection of previously published *Coccidioides* isolates. By providing an open-access standardized tool, cocci-call aims to facilitate cross-study comparisons and advance understanding of *Coccidioides* genomic variation, supporting both clinical and research applications.

## METHODS

### Developing a variant identification pipeline

We developed and benchmarked a variant identification pipeline, *cocci-call*, to (a) assign *Coccidioides* species and (b) identify genome-wide variants. It is publicly available on Github (https://github.com/ksw9/cocci-call). The pipeline was built using the Nextflow workflow language to allow tasks parallelization on High Performance Computing (HPC) clusters.

Moreover, cocci-call employs Docker containers to enhanced reproducibility and ease of deployment. cocci-call comprises two separate workflows: 1) pipeline resources preparation; 2) variant identification. The former workflow makes it easy to automatically download all the required resources, including reference FASTA files (with optional masking steps using RepeatMasker (https://www.repeatmasker.org/) and/or NUCmer (8), BWA indexing (Li & Durbin, 2009), GATK dictionary generation (10), SnpEff database setup (11), and the parallelized download of reference genomes for Kraken2 database generation (12).

Once the necessary pipeline resources have been setup, the variant identification workflow can be run. First, raw reads are preprocessed by trimming low-quality bases (Phred-scaled base quality < 20) and removing adapters with Trim Galore v. 0.6.10 (stringency=1) (https://github.com/FelixKrueger/TrimGalore). This step also generates quality control reports using FastQC (https://www.bioinformatics.babraham.ac.uk/projects/fastqc/). To exclude contamination from other species, a potential source of false genetic variation, Kraken2 (12) is used to taxonomically classify reads and remove those not assigned to the *Coccidioides* genus. A log2-transformed ratio of Kraken2 unique minimizers assigned to *C. immitis* or *C. posadasii* is then used for species assignment according to a user-defined threshold. Regardless of whether the species is assigned to *C. immitis* or *C. posadasii*, all *Coccidioides* reads are retained for downstream analysis, as hybridization between the two sister species has been documented and is a potential source of true biological variation (13). Additionally, users can decide to keep reads not assigned to the *Coccidioides* genus. Next, reads are mapped with bwa v. 0.7.17 (bwa mem) (9) to two reference genomes: GCF_000149335.2 for *C. immitis* and GCA_018416015.2 *C. posadasii*. Duplicate reads are also marked using GATK 4.3 (10) at the end of this alignment step. For samples where the log2-transformed ratio of Kraken2 unique minimizers is below the user-defined threshold, reads are mapped to both genomes, then species is assigned based on the highest mapping percentage. Variants are called with GATK 4.3 HaplotypeCaller and GenotypeGVCFs, setting sample ploidy to 1. Optionally, users can decide to include or exclude non-variant sites in the output vcf. Also, a variant calling step using LoFreq (14) can also be ran separate from GATK. After variant calling, vcf files are filtered using a Python script with customizable filters. By default, SNPs with minimum depth 10X and a minimum variant quality score 20 are retained. This filter also allows the removal of variants with depth or coverage deviating more than two standard deviations from the mean depth and coverage which tend to occur in repeated regions when using unmasked reference genomes. Lastly, filtered variants are annotated using SnpEff (11) and a consensus FASTA sequence is constructed using bcftools 1.17(15) consensus.

### Benchmarking datasets

We evaluated the performance of variant identification in *Coccidioides* in two datasets for which the genomic “truth” is available: a) *in silico* generated short reads from the *C. posadasii* and *C. immitis* reference genomes and b) empirically sequenced *C. posadasii* Δcts2/Δard1/Δcts3 strain NR-166 (BEI) (https://www.beiresources.org/Catalog/Fungi/NR-166.aspx). The reference genomes GCA_000151335.1 (*C. posadasii)* and GCA_004115165.2 (*C. immitis)* were used to simulate short read sequence data using ART (16), generating 10 read sets for each species, with a median coverage of 100X and HS25 model of sequencing error. Note that the selected genomes are different from the ones used by the cocci-call pipeline (GCF_000149335.2 and GCA_018416015.2 for *C. immitis* and *C. posadasii*, respectively), thus making it possible to define “true” variants with respect to the reference. To include an empirically generated benchmarking dataset, we also sequenced the *C. posadasii* Δcts2/Δard1/Δcts3 reference strain NR-166. We extracted DNA with the QIAamp DNA Mini Kit, performed library prep with the NEBNext Ultra II FS DNA Library Prep kit, and sequenced on a single lane of a NovaSeq X Series 10B (150×150 bp) at the University of Utah High-Throughput Genomics (HTG) Shared Resource.

### Generation of a truth SNP set

To generate “truth” VCF files, NUCmer (8) was used to pairwise align assembled genomes used for variant calling to those used for *in silico* sample generation with ART or the reference for the NR-166 strain. NUCmer commands were run with the following parameters: “--maxmatch -- mincluster 100” for *NUCmer*, “-g” for *delta-filter*, default parameters for show-coords, and “-C - I -r -T” for show-snps.

### Measuring performance in alternative benchmarking regions

To evaluate the performance of our pipeline, we identified SNP variants with both cocci-call and MycoSNP (7), using the same set of *in silico* generated short read samples. We then benchmarked small variant calls with <u>vcfdist</u> (Dunn & Narayanasamy, 2023) against the “truth” VCF files described above. For well-characterized species, genomic analysis frequently focuses on standardized benchmarking regions, which often exclude repetitive regions(17) . To identify regions of the genome to exclude from analysis, we applied two tools that identify genomic repeats: RepeatMasker (https://www.repeatmasker.org/) and NUCmer (8).

We then evaluated performance in alternative genomic regions (i.e. the full genome, gene regions only, and regions masked or not masked by RepeatMasker and/or NUCmer) to make recommendations about specific, low-confidence regions that should be excluded from routine *Coccidioides* genomic studies. For each region, we calculated the number of true positives, false positives, true negatives, and false negatives to evaluate model performance using precision, recall, and the F1 score. Precision was defined as the proportion of true positives among all predicted positives, while recall represented the proportion of true positives among all actual positives. The F1 score, calculated as the harmonic mean of precision and recall, provided a single metric to balance false positives and false negatives. This approach simplifies evaluation by combining precision and recall into a single performance metric. The Jaccard index was also calculated for each region to assess the overlap between SNPs detected by cocci-call and MycoSNP, providing a measure of concordance across these regions.

## RESULTS

### User Interaction and Output Features of cocci-call

Users interact with cocci-call by providing a list of short sequence read data and adjusting variant identification parameters to suit their needs. The pipeline’s output includes sample-specific VCF files, a summary file with species assignments, and sample coverage and quality metrics. A key feature of cocci-call is its species assignment method, which leverages a log-transformed Kraken2 minimizer ratio. This ratio, representing the proportion of subsequences uniquely classified to each species, allows for a customizable threshold to ensure accurate and flexible assignments based on user requirements.

### Species identification performance

cocci-call assigned *Coccidioides* species in 20 *in silico* read sets (10 from *C. immitis* and 10 from *C. posadasii)* with 100% accuracy (Fig. 2). We additionally tested the performance of cocci-call in identifying species in publicly available sequence data from previous studies, using the species assigned by the submitting study as the gold standard. Among these empirically collected isolates, cocci-call assigned species with 98.86% (173 of 175) accuracy (Fig. 3). A single sample, which was labeled unknown by the submitting study, was not definitively assigned to a species, but had a log-transformed *immitis*/*posadasii* Kraken2 minimizers ratio of 1.12, suggesting it was likely *C. immitis* or a *C. immitis* strain with introgressed regions (Fig. 3). In another sample, cocci-call assigned *C. posadasii* though the submitting study assigned the sample *C. immitis;* this was potentially an instance of misassignment in the original study (Fig. 3).

**Fig. 1.**
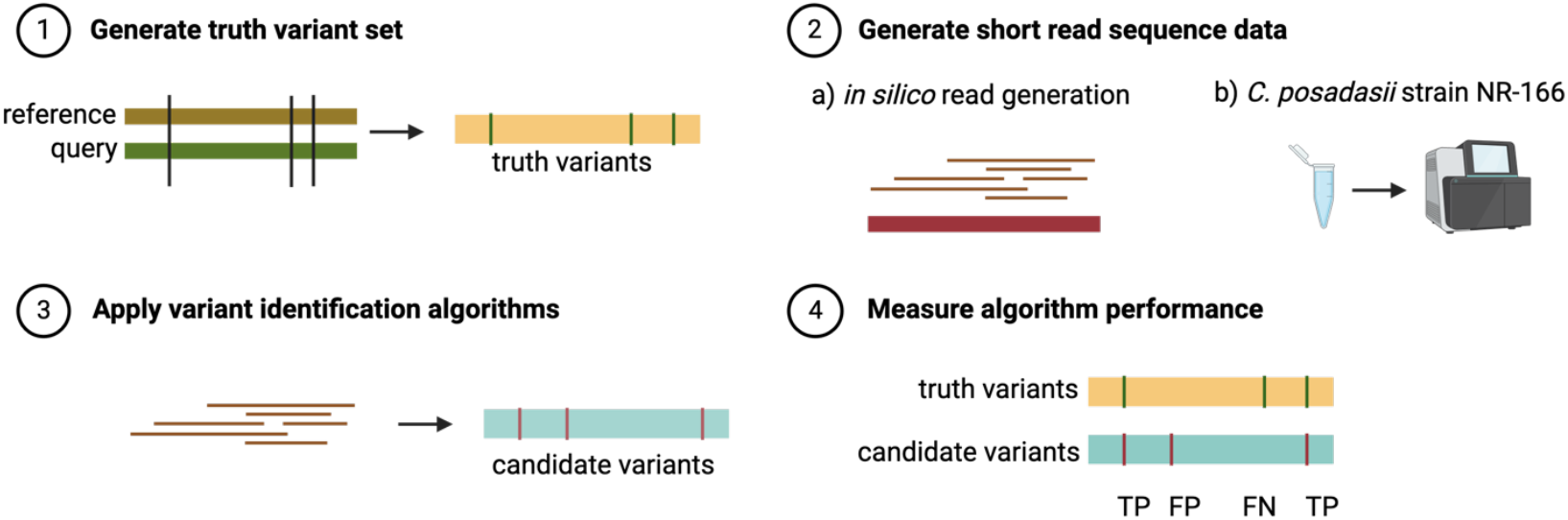
Workflow for benchmarking variant identification algorithms in *Coccidioides* species. Step 1: Generate truth variant set. The true set of genetic variants is identified by comparing a reference genome with a query genome. This comparison produces a set of “truth variants” that serve as a benchmark. Step 2: Generate short-read sequence data. Short-read sequence data is generated in two ways: (a) *in silico* read generation, where simulated reads are computationally generated from the query genome; and (b) sequencing of *C. posadasii* strain NR-166, where biological reads are obtained experimentally. Step 3: Apply variant identification algorithms. The generated sequence reads are processed through variant-calling algorithms, producing a set of “candidate variants” that can be compared to the truth variant set. Step 4: Measure algorithm performance. The accuracy of the variant-calling algorithm is assessed by comparing the truth variant set to the candidate variants, with performance metrics calculated based on true positives (TP), false positi ves (FP), and false negatives (FN).

**Fig. 2.**
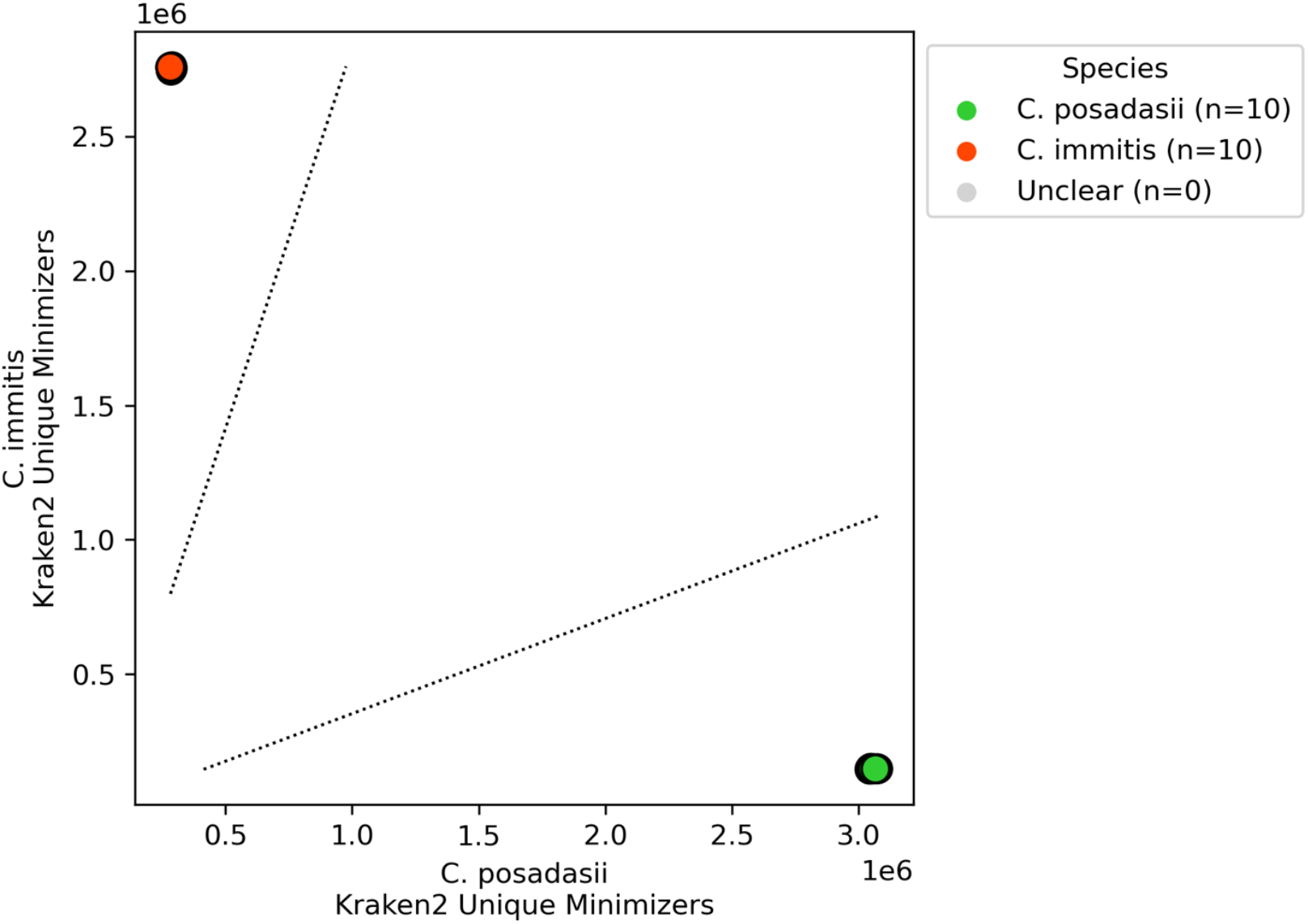
Accuracy of *Coccidioides* species assignment by cocci-call for *in silico* sequence data. Points indicate unique sequence data generated *in silico* from reference genomes of both *C. immitis* and *C. posadasii* (indicated by point color). Points are located along the x-axis by the number of k-mers that uniquely map to the *C. posadasii* reference genome and along the y-axis by the number of k-mers that uniquely map to the *C. immitis* reference genome. The dashed line denotes the species assignment threshold, demonstrating cocci-call’s ability to distinguish between *C. posadasii* and *C. immitis*. Gray circles (n=0) would represent samples with unclear species assignment, though none were observed in this dataset.

**Fig. 3.**
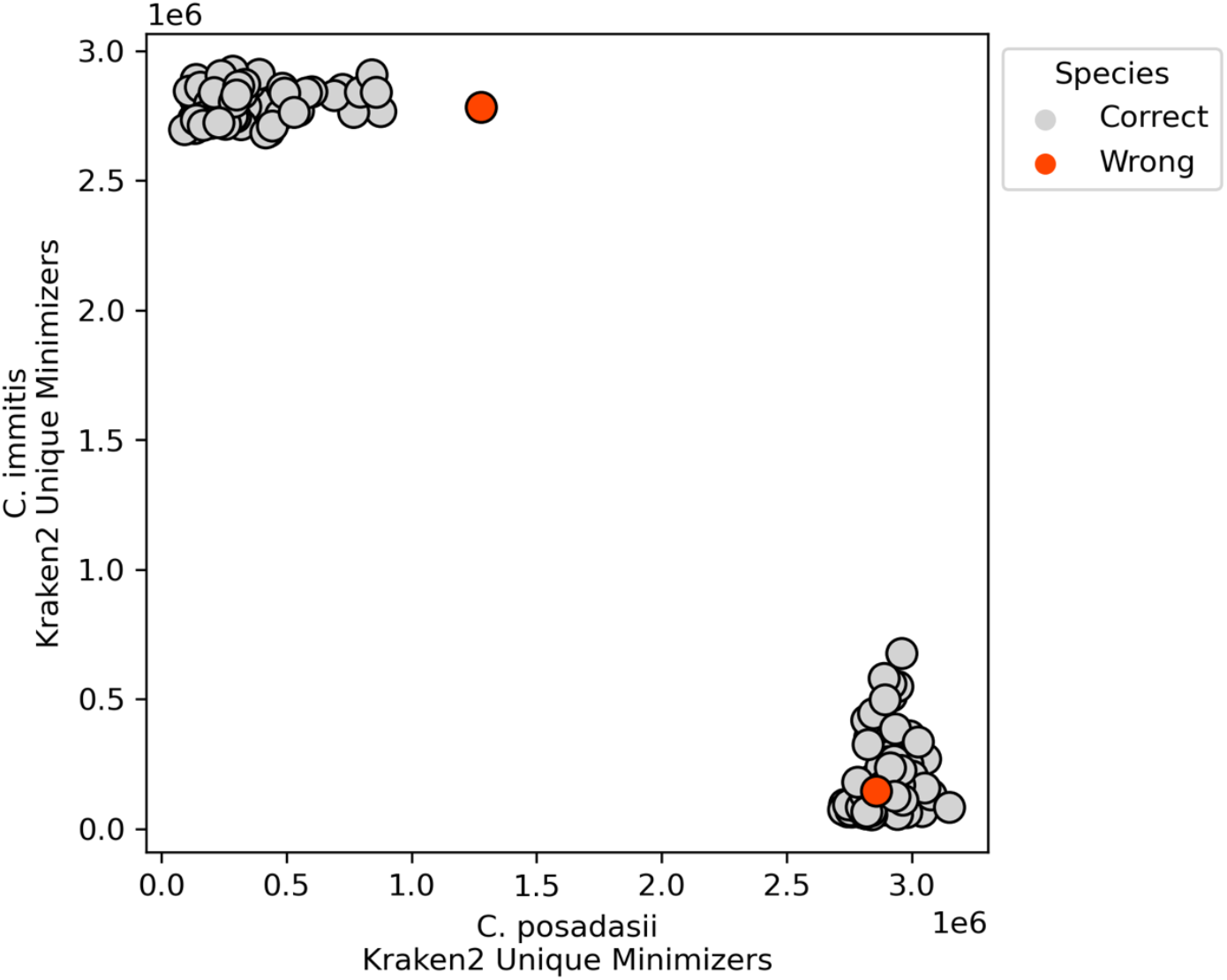
Accuracy of *Coccidioides* species assignment by cocci-call for previously published *Coccidioides* sequence data. Gray circles represent correctly assigned samples, with clusters corresponding to C. posadasii and C. immitis based on Kraken2 minimizer values along the horizontal and vertical axes, respectively. The orange circles indicate misclassified samples.

### Variant identification performance prior to masking

We assessed SNP detection using the variant-calling tools cocci-call and MycoSNP, on *in silico* whole-genome datasets for *C. immitis* and *C. posadasii*. In the *C. immitis* datasets, cocci-call identified a median of 78,182 SNPs (IQR: 78,104–78,334), while MycoSNP detected a median of 38,065 SNPs (IQR: 38,053–32,111) across 10 simulated datasets (Figure 4A). Among these, 32,067 of the SNPs were consistently detected by both pipelines, highlighting a core overlap that underscores shared detection capabilities. This overlap constitutes approximately 41% of the SNPs identified by cocci-call and 84% of those detected by MycoSNP. The Jaccard index of 0.34 for *C. immitis* indicates a moderate level of concordance between cocci-call and MycoSNP..

**Fig. 4.**
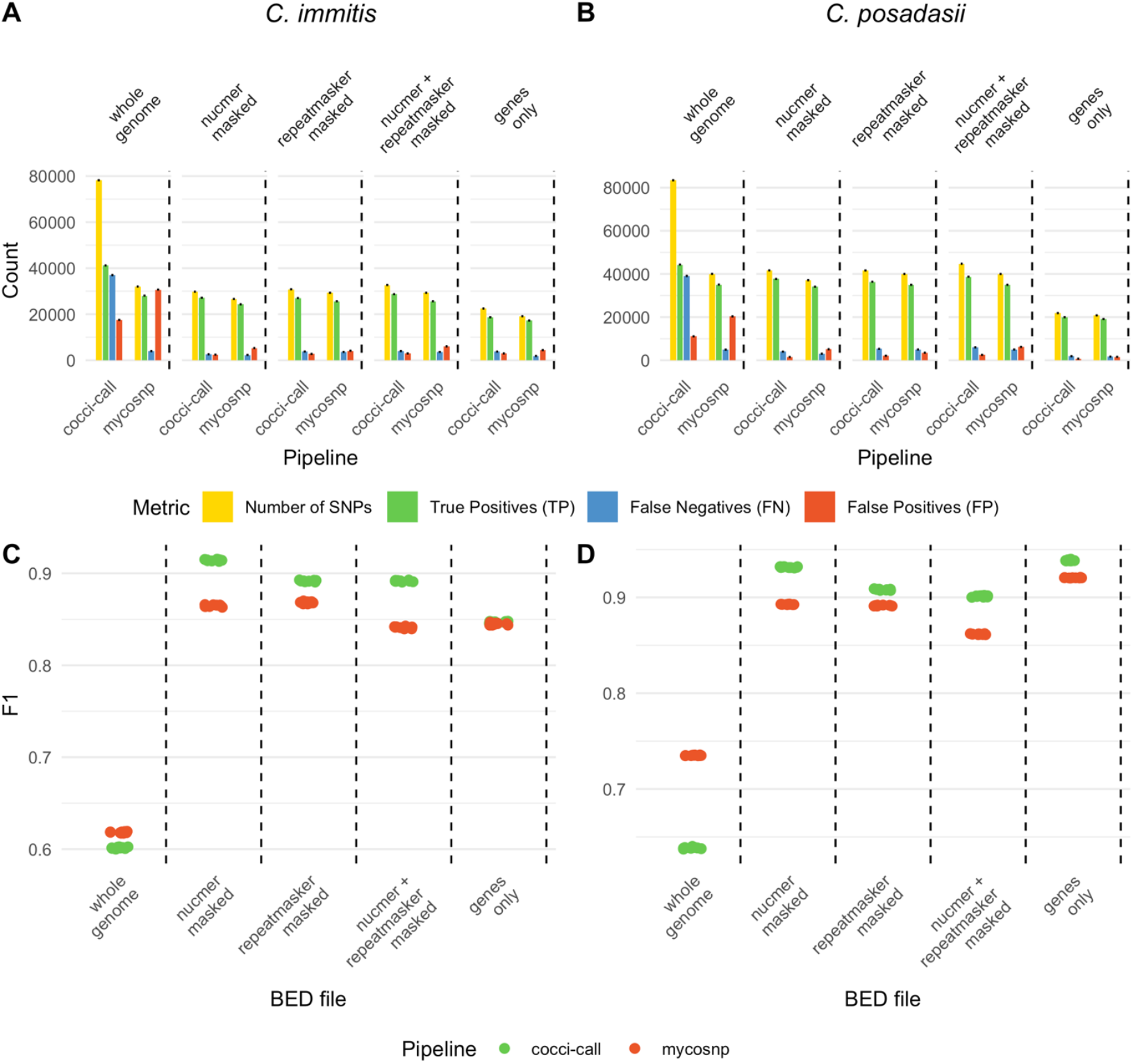
Magnitude of *Coccidioides* variants identified and performance across benchmarking regions for *in silico* sequence data. Total SNPs (yellow), true positives (TP, green), false negatives (FN, blue), and false positives (FP, red) identified by the cocci-call and MycoSNP pipelines for (A) *C. immitis* and (B) *C. posadasii*. Vertical lines delimit different benchmarking regions: the whole genome, NUCmer-masked, RepeatMasker-masked, combined NUCmer and RepeatMasker-masked, and gene-only regions. Panels (C) and (D) show the F1 scores of the cocci-call (green) and MycoSNP (red) pipelines across the same benchmarking regions.

Both *cocci-call* and MycoSNP achieved similar F1 scores on the unmasked genomes, pointing to comparable accuracy (Fig. 4C). Yet, a detailed examination reveals contrasting strengths: cocci-call demonstrated higher sensitivity (70.1%, IQR: 70.0%–70.1%), capturing a larger proportion of true positive variants, whereas MycoSNP showed significantly higher precision (87.5%, IQR: 87.5%–87.6%), indicating fewer false positives (Fig 4B; S1A–B). The resulting F1 scores were 60.1% (IQR: 60.0%–60.2%) for cocci-call and 61.8% (IQR: 61.7%– 61.8%) for MycoSNP (Fig 4B).

In *C. posadasii*, the trends were similar. Here, cocci-call identified a median of 83,400 SNPs, while MycoSNP detected 40,018 SNPs (Fig. 4B). Of the total SNPs identified, 40,163 were detected by both cocci-call and MycoSNP. This accounts for 38.9% of the SNPs identified by cocci-call and 79.7% of those detected by MycoSNP, with a Jaccard index of 0.36 for *C. posadasii*, indicating a moderate level of agreement between the two pipelines for this species.

Further analysis of *C. posadasii* variant calling revealed differences in tool performance. While MycoSNP attained a higher F1 score (73.4%, IQR: 73.4%–73.5%) compared to cocci-call’s 63.8% (IQR: 63.8%–63.8%), cocci-call demonstrated a notably higher sensitivity, detecting 79.9% (IQR: 79.9%–80.0%) of true variants versus MycoSNP’s 63.3% (IQR: 63.2%– 63.3%) (Fig. 4D; S1C). However, MycoSNP outperformed cocci-call in precision, achieving 87.6% (IQR: 87.5%–87.6%) compared to cocci-call’s 53.1% (IQR: 53.0%–53.1%) (Fig. S1D).

### Choice of benchmarking region impacts performance of *Coccidioides* variant identification

We compared genomic regions identified by two commonly used tools for repeat identification. Overall, both NUCmer and RepeatMasker identify similar repetitive regions in both species (Fig. S2). However, certain contigs (e.g., *C. immitis*: NW_004504306.1; *C. posadasii*: CP075073.1 and CP075074.1) showed significantly less overlap in repetitive regions identified by the two tools, with RepeatMasker identifying most of the repetitive regions in these cases. This discrepancy highlights variability in repetitive region detection across specific genomic regions and underscores the importance of using multiple tools to achieve comprehensive coverage.

The selection of benchmarking regions significantly impacted the performance of both variant-calling pipelines. Focusing the analysis on gene-only regions or masking repetitive regions led to substantially higher F1 scores compared to genome-wide analysis. This improvement highlights the tradeoff between accuracy and variant yield: while genome-wide approaches capture more total SNPs, they introduce noise from repetitive sequences that result in false variant calls. In contrast, applying stringent masks or restricting to high-confidence regions enhances accuracy by reducing false positives, but at the cost of fewer variants for downstream analysis (Fig. 4).

For *C. immitis*, cocci-call’s F1 score increased from 60.2% (IQR: 60.1%–60.2%) to 84.7% (IQR: 84.6%–84.8%) when analysis was limited to gene-only regions (Fig 4B). Similarly, MycoSNP’s F1 score rose from 61.9% (IQR: 61.8%–61.9%) to 84.5% (IQR: 84.4%–84.6%) under the same conditions (Fig 4B). Masking repetitive regions using NUCmer further elevated performance, with cocci-call achieving an F1 score of 91.4% (IQR: 91.4%–91.5%) and MycoSNP reaching 86.5% (IQR: 86.4%–86.5%) (Fig 4B). When additional filtering was applied using RepeatMasker, alone or combined with NUCmer, similar trends emerged (Fig 4B).

cocci-call and MycoSNP were largely consistent in SNP identification. In gene-only regions, for example, 71% of total unique SNPs overlapped, encompassing 80% of cocci-call and 87% of MycoSNP SNPs, resulting in a Jaccard index of 0.71. RepeatMasker-masked regions showed a stronger concordance, with 78% of total unique SNPs overlapping, covering 93% of cocci-call and 83% of MycoSNP SNPs, yielding a Jaccard index of 0.78. NUCmer-masked regions exhibited a similarly high overlap, with 79% of unique SNPs shared, covering 88% of cocci-call and 89% of MycoSNP SNPs, reflected in a Jaccard index of 0.79. The masked union, combining RepeatMasker and NUCmer regions, demonstrated a SNP overlap of 74%, representing 88% of cocci-call and 83% of MycoSNP SNPs, with a Jaccard index of 0.74.

Similar to *C. immitis*, restricting the analysis to gene-only regions in *C. posadasii* resulted in a significant improvement, with cocci-call’s F1 score rising markedly from 63.9% (IQR: 63.8%– 63.9%) to 93.8% (IQR: 93.8%–93.9%) (Fig. 4D). MycoSNP’s F1 increased from 73.5% (IQR: 73.5%–73.5%) to 92.0% (IQR: 92.0%–92.1%) (Fig 4D). Similarly to *C. immitis*, masking repetitive regions using NUCmer produced considerable gains, with cocci-call achieving an F1 score of 93.1% (IQR: 93.1%–93.2%) and MycoSNP reaching 89.3% (IQR: 89.3%–89.3%). When further filtering was applied using RepeatMasker, cocci-call maintained high accuracy with an F1 score of 90.8% (IQR: 90.8%–90.8%), while MycoSNP scored 89.1% (IQR: 89.1%– 89.2%) (Fig 4D). Combining NUCmer and RepeatMasker also proved beneficial, with cocci-call achieving 90.1% (IQR: 90.1%–90.2%) and MycoSNP reaching 86.2% (IQR: 86.1%–86.2%) (Fig 4D).

In gene-only regions, cocci-call and MycoSNP demonstrated a high level of concordance, with approximately 84% of cocci-call SNPs and 90% of MycoSNP SNPs overlapping, leading to a Jaccard index of 0.84. RepeatMasker-masked regions showed similarly robust agreement, with 75% of total unique SNPs overlapping, representing 92% of cocci-call and 80% of MycoSNP SNPs, and a Jaccard index of 0.75. NUCmer-masked regions exhibited an even higher overlap, with 79% of total unique SNPs shared, covering 87% of cocci-call and 89% of MycoSNP SNPs, corresponding to a Jaccard index of 0.79. The intersection of RepeatMasker and NUCmer-masked regions reflected high concordance, with 71% of unique SNPs overlapping, accounting for 86% of cocci-call and 80% of MycoSNP SNPs, yielding a Jaccard index of 0.71.

### Variant identification performance in empirically generated sequence data

We measured genome-wide performance on sequence data from *C. posadasii* strain NR-166 using F1 scores, with cocci-call achieving 63.9% (IQR: 63.8%–63.9%) and MycoSNP 73.5% (IQR: 73.5%–73.5%). Focusing on gene-only regions, cocci-call’s F1 score improved significantly to 93.8% (IQR: 93.8%–93.9%), while MycoSNP reached 92.0% (IQR: 92.0%– 92.1%) (Fig. 5). Masking repetitive regions with NUCmer further enhanced cocci-call’s F1 to 93.1% (IQR: 93.1%–93.2%) and MycoSNP’s to 89.3% (IQR: 89.3%–89.3%) (Fig. 5). Even when using RepeatMasker or a combined approach with both NUCmer and RepeatMasker, cocci-call continued to perform better than MycoSNP, although both pipelines experienced slight F1 decreases in masked repetitive regions (Fig. 5).

**Fig. 5.**
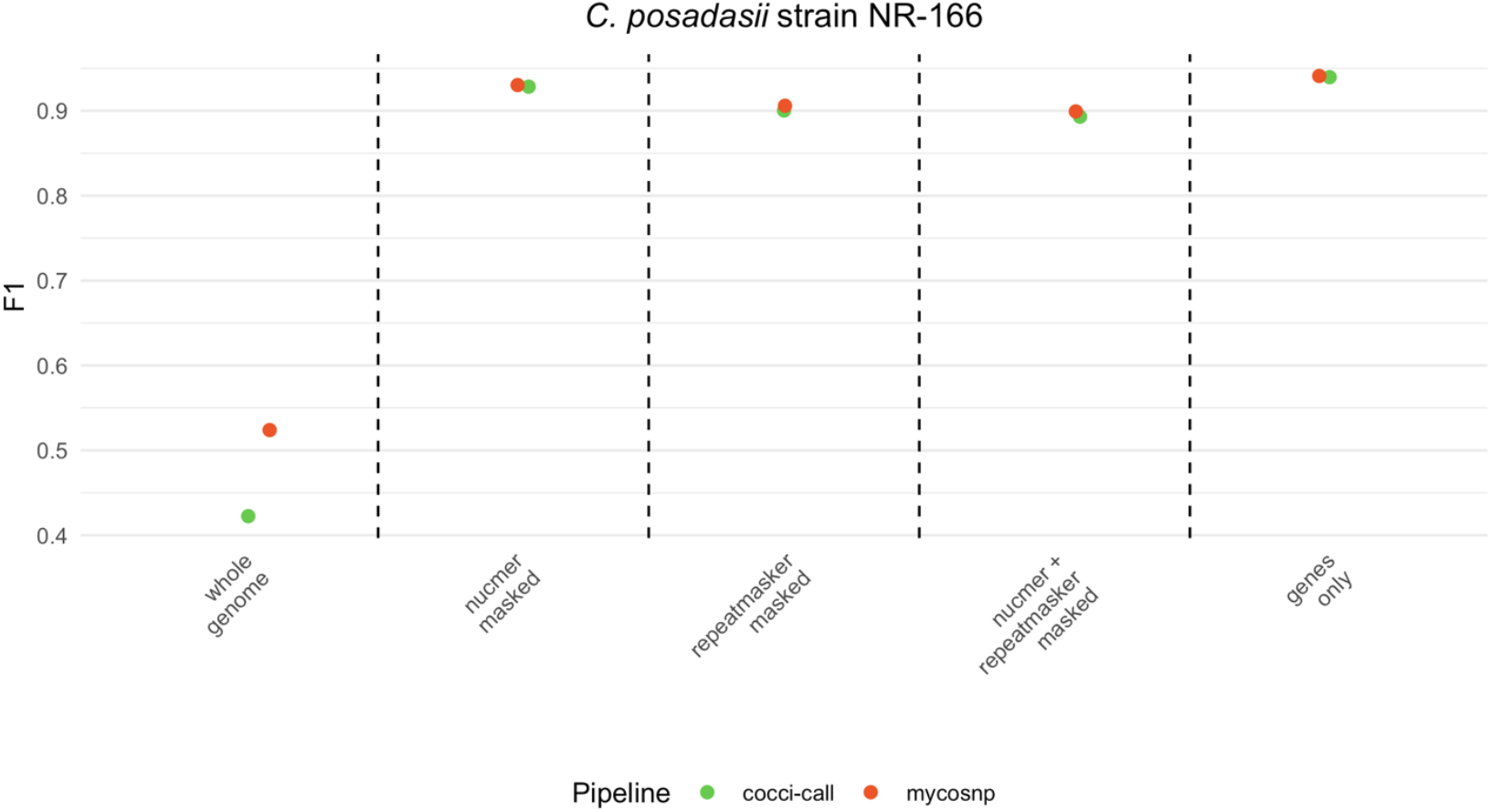
Performance of variant identification across benchmarking regions for empirically generated sequence data. Comparison of F1 scores for variant detection by cocci-call (green) and MycoSNP (red) across different genomic regions in *C. posadasii* strain NR-166. Vertical lines delimit different benchmarking regions: the whole genome, NUCmer-masked, RepeatMasker-masked, combined NUCmer and RepeatMasker-masked, and gene-only regions.

## DISCUSSION

Here, we introduce cocci-call, an open-source pipeline developed to assign species and identify variants in the emerging fungal pathogens *C. immitis* and *C. posadasii*. cocci-call performs well both in assigning species in *in silico* and empirically generated sequence data and in recovering known SNP variation. Importantly, performance of variant identification improves dramatically when focusing on high confidence benchmarking genomic regions.

Accurate species identification and variant detection are critical for public health interventions, such as outbreak monitoring and infection source tracking (18,19). The integration of Kraken2-based taxonomic filtering in cocci-call not only enables accurate species identification but also improves variant detection by removing non-target reads. This dual functionality helps reduce false positives, particularly in complex multi-organism samples like clinical and environmental datasets. By focusing exclusively on *Coccidioides* spp. reads, this approach enhances the accuracy and biological relevance of identified variants, offering significant benefits for both research and clinical applications.

Species with well-characterized genomes often have standard benchmarking regions— specific genomic regions where the performance of variant identification has been systematically evaluated (e.g., Bagal et al., 2022; Majidian et al., 2023). This study is the first to assess the impact of benchmarking region choice on variant identification in *Coccidioides*. Our findings reveal that the choice of benchmarking region not only significantly influences the accuracy of variant identification but also affects the total number of true SNP variants identified. This tradeoff between variant detection accuracy and the magnitude of genomic variation available for downstream analyses underscores the importance of benchmarking region selection. To address the diverse requirements of users, the cocci-call pipeline includes an adjustable parameter for selecting benchmarking regions. This flexibility allows users to customize their analyses according to specific research goals and datasets.

cocci-call’s per-sample approach contrasts with MycoSNP’s reliance on joint variant calling. This design enables cocci-call to maintain comparable precision while achieving higher sensitivity, particularly in datasets with limited population diversity. This sensitivity is advantageous for studies involving individual-level data or sparse sample sets, allowing for the detection of low-frequency variants critical for diagnostics and surveillance. While MycoSNP benefits from its joint-calling strategy, which can boost precision in population studies, cocci-call’s flexibility broadens its applicability across diverse research and clinical contexts.

Our findings emphasize that both cocci-call and MycoSNP (7) are high-performance tools, each demonstrating strong capabilities in variant detection. Both pipelines achieve comparable overall performance based on F1 scores, highlighting their effectiveness in addressing the challenges of *Coccidioides* genomic data. However, the choice of benchmarking region significantly impacts performance metrics, with cocci-call excelling in gene-only and filtered repetitive regions, achieving F1 scores exceeding 84%. For empirical datasets, cocci-call maintains consistent robustness, with F1 scores surpassing 93% in filtered regions, addressing limitations seen in repetitive areas. These complementary strengths underscore the value of cocci-call as a tool for standardizing species assignment and variant identification for *Coccidioides* spp., particularly in clinical and epidemiological applications.

While cocci-call demonstrates robust performance, we identify limitations. One limitation is its reliance on short-read sequencing, which may not accurately recover variation in repetitive regions. Integrating long-read sequencing technologies, like Nanopore or PacBio, could improve variant detection in these challenging genomic areas. Additionally, incorporating joint variant calling capabilities would expand its applicability. At present, the pipeline is optimized for SNP detection, given their central role as a primary source of genetic variation used in phylogenetic inference. Future efforts will focus on benchmarking cocci-call for other types of genomic variation, including insertions and deletions, structural variants, and copy number variations. Expanding its capabilities to accommodate these additional variant classes will not only broaden its scope but also strengthen its utility in addressing diverse research questions in epidemiology and population genomics.

Our results demonstrate cocci-call’s potential as a validated tool for both research and clinical applications. In endemic regions, accurate differentiation between *C. immitis* and *C. posadasii* is essential for tracking infections, managing outbreaks, and understanding pathogen dynamics (3,21). cocci-call’s accurate species assignment and variant detection capabilities make it a valuable tool for infection tracking, including cases associated with travel or migration (22). Finally, cocci-call is a tool for standardized and reproducible genomic epidemiology, enabling cross-study comparisons, advancing *Coccidioides* genomics, and fostering collaboration among research groups.

## Data Availability

Raw sequencing data generated for this study are available in the Sequence Read Archive (SRA) under BioSample SAMN45887705. This includes whole-genome sequencing data used for variant identification and benchmarking, as well as sequencing data from clinical and environmental Coccidioides samples. The pipeline cocci-call, developed for species-aware variant identification in Coccidioides spp., is publicly available at https://github.com/ksw9/cocci-call. All scripts and code used for the analysis performed in this study are included in the pipeline repository.

## ABBREVIATION

BEI: Biodefense and Emerging Infections Research Resources Repository
CDC: Centers for Disease Control and Prevention
FN: False Negative
FP: False Positive
F1 Score: Harmonic mean of precision and recall
GATK: Genome Analysis Toolkit
IQR: Interquartile Range
NEB: New England Biolabs
NUCmer: Nucleotide MUMmer
SNP: Single-Nucleotide Polymorphism
TP: True Positive
VCF: Variant Call Format
WGS: Whole-Genome Sequencing.

## AUTHOR CONTRIBUTIONS

M.M. and K.S.W. designed the project and conducted the analysis. All authors contributed to interpreting the results and engaged in in-depth discussions. E.M.F. took the lead in writing the manuscript, with support from all co-authors. The final manuscript was reviewed, edited, and approved by all authors.

