## Supporting Information for "cocci-call: a species-aware variant identification pipeline for *Coccidioides* spp. in genomic epidemiology"

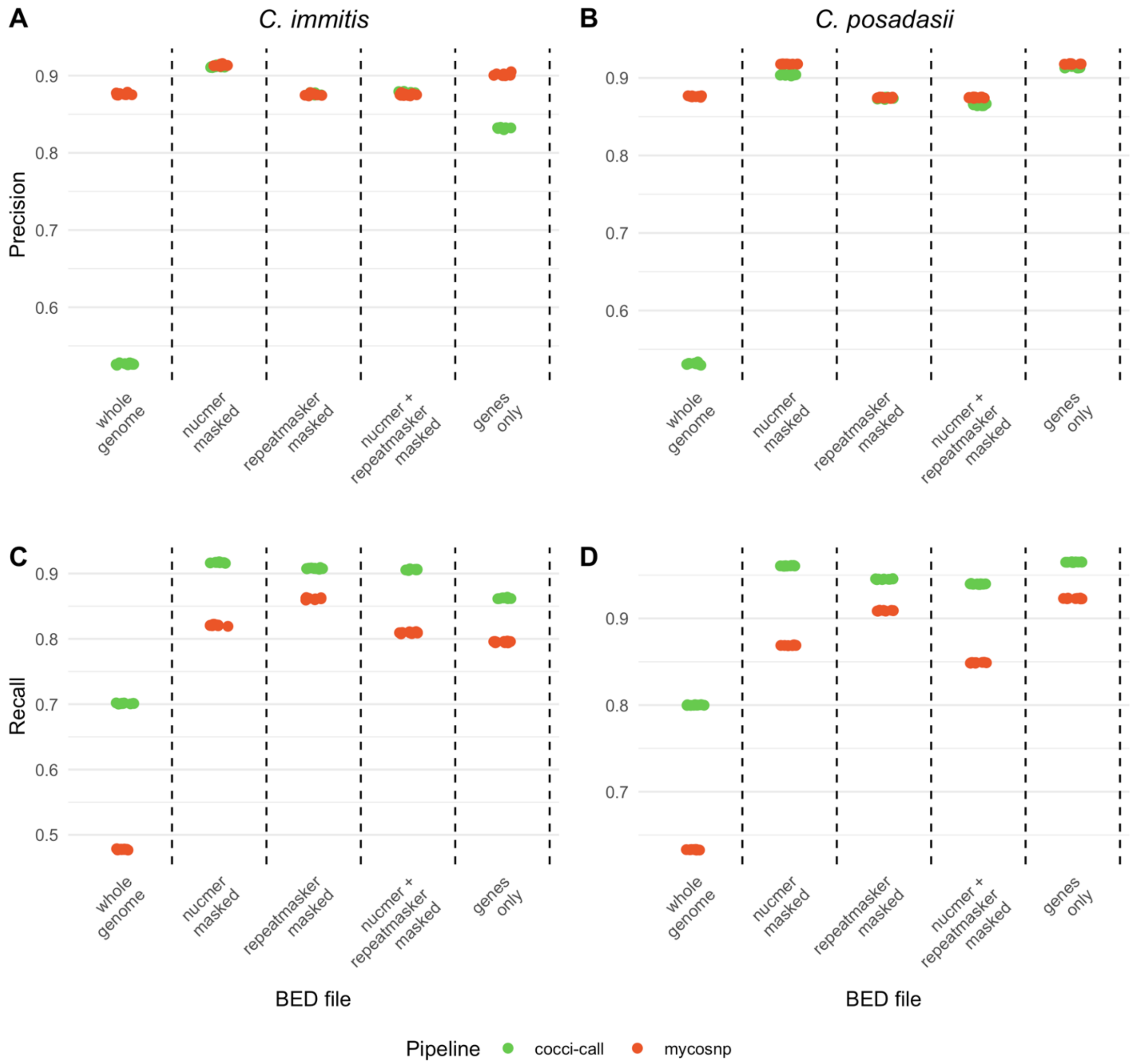


**Fig. S1. Comparison of precision and recall for *Coccidioides* variant identification**. (A,B) Precision and (C,D) recall of variants identified by cocci-call (green) and MycoSNP (orange). Horizontal facets indicate benchmarking region: the whole genome, NUCmer-masked, RepeatMasker-masked, combined NUCmer and RepeatMasker-masked, and gene-only regions.

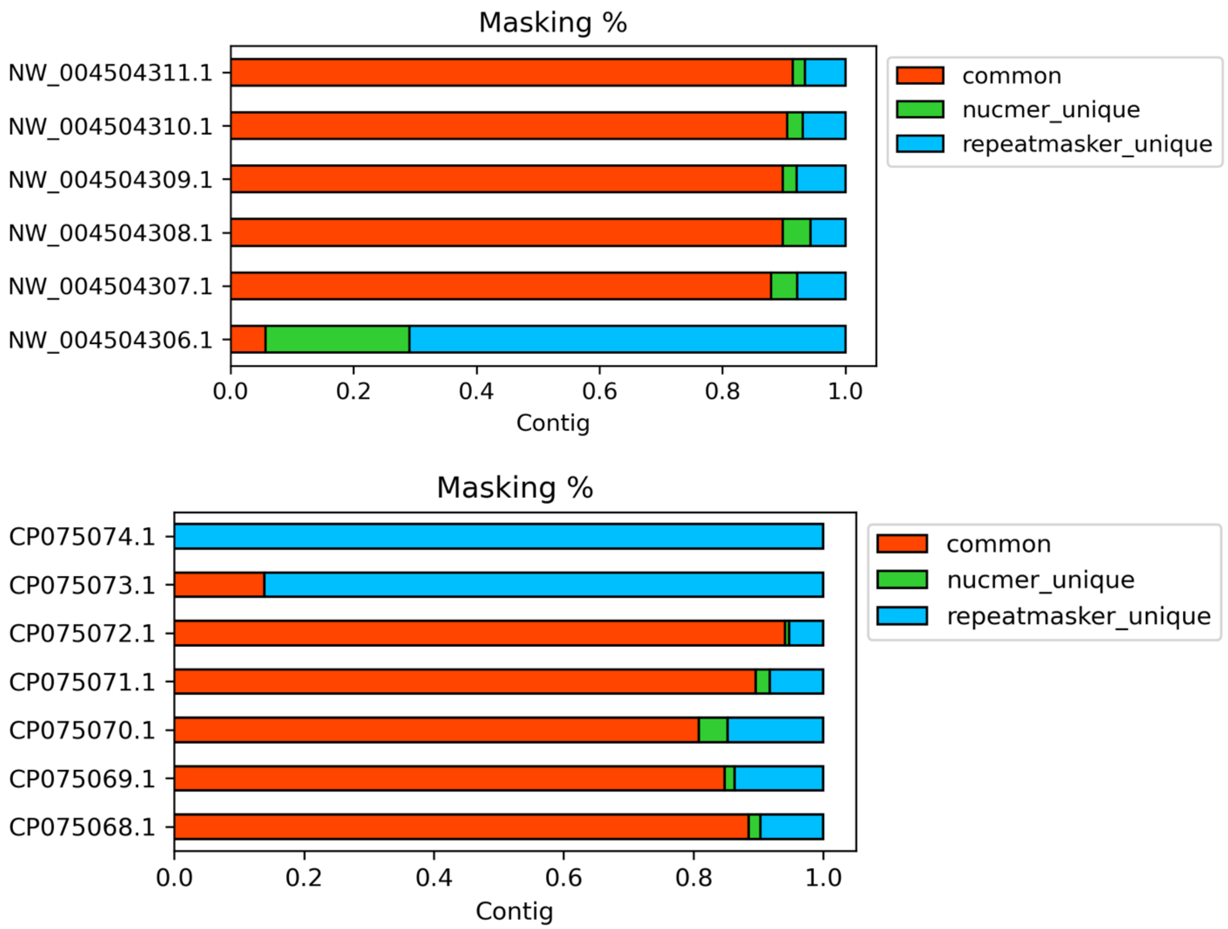


**Fig. S2. Repetitive genomic regions identified by different tools.** Bars indicate the percentage of the contig identified as repetitive for *C. immitis* (top) and *C. posadasii* (bottom). Colors indicate the types of masking: "common" regions identified by both NUCmer and RepeatMasker (shown in red), regions unique to NUCmer ("NUCmer_unique," shown in green), and regions unique to RepeatMasker ("repeatmasker_unique," shown in blue).
